# Exploring the Continental Staff System as a Framework for the Hospital Incident Command – Protocol for a Qualitative Grounded Theory Study

**DOI:** 10.1101/2025.05.01.25326786

**Authors:** M von der Forst, E Popp, H Schaefer, K Bikowski, M Ries, J Kuellenberg

**Author notes:** **Corresponding Author:** Maik von der Forst, MD, MHBA, DESAIC, Crisis and Disaster Management Unit, University Hospital Heidelberg, Im Neuenheimer Feld 420, 69120 Heidelberg, Germany.

## Abstract

**Background:** In order to maintain hospital functionality during exceptional situations, the hospital incident command must be adequately empowered and prepared. As hospitals in Germany and elsewhere are legally required to implement a Hospital Emergency and Response Plan, regular training exercises are essential to ensure that staff acquire the necessary knowledge and competencies.

**Objectives:** This study will investigate the transition process from routine clinical operations to an exceptional situation during the training of a continental staff-system based hospital incident command. Primary objective: 1) deepen the understanding of leadership communication and dynamics, 2) assess training outcomes for hospital incident command teams. Secondary objectives: examine a. the applicability of the traditional incident command model in hospitals with limited staff experience, b. evaluate participants’ operational confidence, c. analyze communication within command functions, and d. identify the strengths and weaknesses of a structured hospital incident command system

**Methods:** The study is based on a staff framework exercise conducted following targeted training. Data will be gathered prospectively through semi-structured interviews with participants, and an analysis of observational data, including notes and protocols recorded during the exercise. All data will be analyzed following Grounded Theory methodology, a systematic approach to theory construction through qualitative data analysis.

**Discussion:** Despite the growing importance of hospital incident command systems in disaster and business continuity management, research in this area remains limited. This study aims to fill the gap by examining the implementation and functioning of a continental staff system-based command in hospitals, with a particular focus on transition dynamics and internal communication processes.

## Introduction

In disaster scenarios, all relevant actors must rapidly coordinate their actions to achieve shared objectives and carry out defined responsibilities. Hospitals in Germany and elsewhere are legally required to maintain a Hospital Emergency and Response Plan which includes an obligation to train staff the requisite knowledge and skills. Regular training exercises in hospital emergency and response planning are therefore strongly recommended (1). In particular, the hospital incident command—the designated leadership body —should be empowered to ensure the continuity of hospital operations during exceptional situations.

The hospital emergency and response handbook by the Federal Office of Civil Protection and Disaster Assistance recommends aligning the hospital command organizational structure with the continental staff system service regulation DV 100 of the Standing Conference on Disaster Prevention and Civil Protection, as well as the operational guideline for non-police emergency response by specialized authorities (2). This structure adopts a staff-based model, i.e., a temporarily established advisory body that supports leadership functions in complex operational environments (p. 31(3,4). Staff work, as a distinct mode of operations in high-risk contexts, is characterized by uncertain duration, diverse task demands, limited information, high time pressure, and restricted opportunities for training (3), similar to so-called VUCA situations (5). Consequently, actors require expertise in emergency protocols, problem-solving, decision-making, and leadership (6). Regular training not only enhances procedural knowledge but also improves role clarity, teamwork, communication, confidence in response capabilities, and stress resilience during actual emergencies (6).

However, scientific literature highlights a lack of sufficient evidence to conclusively determine whether training in healthcare settings effectively improves disaster preparedness (7). Several studies call for a stronger emphasis on the scientific evaluation of the efficacy of such training programs (8), particularly through high-quality multidisciplinary research using validated assessment tools (9). Despite these recommendations, research on staff-based leadership models in hospitals during exceptional situations remains notably limited.

Against this backdrop, investigating leadership communication and dynamics during the transition from routine operations to emergency mode, as well as evaluating the training outcomes for hospital incident command teams, holds significant potential for strengthening hospital crisis management. This study seeks to systematically evaluate these experiences. A deeper understanding of command dynamics may enhance leadership effectiveness and operational readiness, while structured training assessments could refine curricula and address staff-specific needs. Furthermore, identifying barriers to implementation and sources of resistance is essential for the successful adoption of hospital emergency planning. Targeted strategies to overcome these obstacles may ultimately improve compliance, coordination, and overall response effectiveness.

## Objectives

There is a lack of evidence as to which organizational form is functional for the management of special situations in hospitals. This study therefore aims to generate new findings on the use of a staff-based incident command system and to analyze transition and communication processes within hospital operations management (see Table 1).

**Table 1:** Objectives and Methods of the OrAKEL Study.

| Objective Type | Objective Number | Objective focus | Qualitative Assessment Methods |
| --- | --- | --- | --- |
| **Primary** | 1 | Leadership communication and dynamics in the transition from routine clinical operations to an emergency situation. | **Triangulation of Data Sources:**<br>✓ Semi-structured interviews (iteratively refined through theoretical sampling)<br>✓ Participant observation (open, non-structured observations, field notes, photographic documentation)<br>✓ Continuous evaluation of the exercise by the external service provider (pre-briefing, debriefing, feedback, coaching) |
|  | 2 | Training and exercise effects within a structured (according to DV-100) hospital incident command system, including challenges, resistance, benefits, subjective participant experiences, potential improvements, and alternatives. |  |
| **Secondary** | a | Applicability of the traditional incident command concept in hospitals, considering a workforce largely inexperienced in continental staff system-based work. |  |
|  | b | Participants' operational confidence. |  |
|  | c | Communication behaviors within the command functions. |  |
|  | d | Advantages and disadvantages of a structured hospital incident command system. |  |

The primary objectives of the present study are:

1. Exploration of leadership communication and dynamics in the transition process from routine clinical operations to an emergency situation
2. Assessment of training and exercise effects within a structured hospital incident command system, including challenges, resistance, benefits, subjective participant experiences, potential improvements, and alternatives

Additionally, the following secondary objectives will be investigated:

a. Examination of the applicability of the traditional incident command concept in hospitals, considering a workforce largely inexperienced in continental staff system-based work.
b. Evaluation of participants’ operational confidence
c. Analysis of communication behaviors within the command functions.
d. Assessment of the advantages and disadvantages of a structured hospital incident command system

## Methods

The study follows the principles of the spirit statement an overview of the recruitment process and study-specific measures is stated in Figure 1(10). Participant recruitment will start May 1^st^ and will be completed within May 25^th^ before the exercise sessions start and data collection will be completed until December 2025, the results are expected in the first half of 2026. The study is expected to be completed in October 2026. The actual version of the described protocol is V1.1, 03.03.2025, which has also been approved by the Ethics Committee (see Ethics and Dissemination).

**Figure 1:**
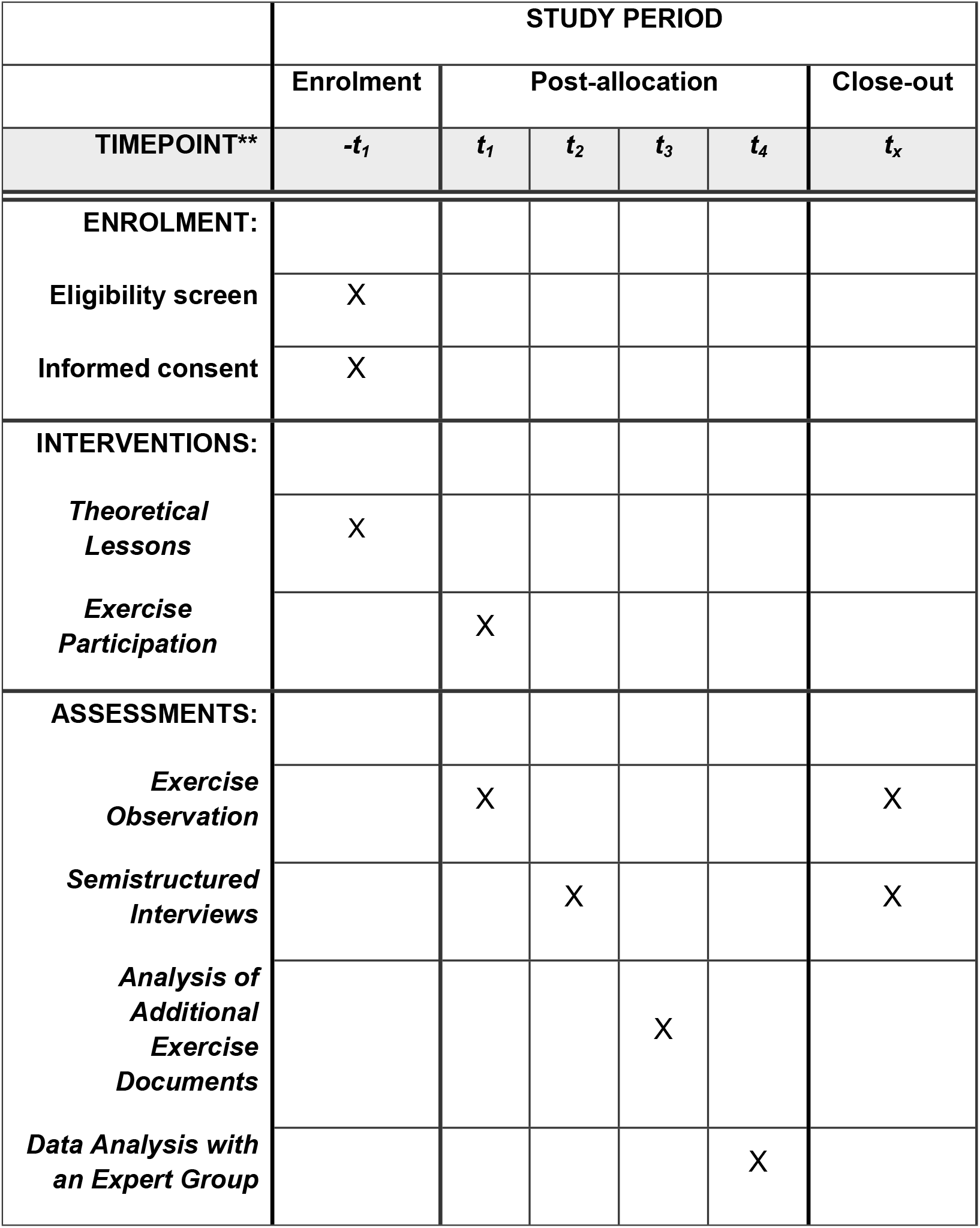
Schedule of Enrolment, Interventions, and Assessments for the OrAKEL Study, following the Spirit Statement(10)

This study is designed as a qualitative, single-site investigation using a constructivist Grounded Theory approach.

### Qualitative Methods

The methodology is based on the Grounded Theory Methodology: a systematic methodology for theory construction through qualitative data analysis. This approach emphasizes the generation of theories directly from empirical data, rendering it particularly valuable for comprehending complex social phenomena (11).

In the Grounded Theory Methodology (GTM), the rule of thumb “All is data” applies, whereby interviews, document analyses (observation sheets, field notes, photo documentation of flipcharts, etc. during the exercise) and observations are included in the study as equally valid data sources. This diversity enables a comprehensively holistic coverage of the research field and supports the gradual development of the theory.

### Sampling (Study Population)

The process of data collection is continued until no new theoretically relevant findings can be obtained - a state known as theoretical saturation. In addition, GTM follows the logic of so-called theoretical sampling, which takes place step by step and is based on the theoretical relevance of the cases. Instead of a one-off data collection with subsequent evaluation, data is collected and analyzed iteratively. Each analysis phase serves as a basis for the targeted selection of the next case in order to systematically expand theoretical understanding and drive theory development forward in a targeted manner (12).

In this study, the theoretical sample basically includes all employees of Heidelberg University Hospital who could be deployed into the hospital incident command system. Further participation in at least one hospital incident command training course or/and in one exercise as well as written declaration of consent from the employee are necessary. Furthermore, only adult employees who are capable of giving informed consent will be included in the study. The sole exclusion criterion is a refusal to provide informed consent for participation in the study. A written informed consent will be obtained by members of the study team before inclusion. Approximately 20 participants will be recruited, with the final sample size determined based on empirical indicators of theoretical saturation (13).

### Methodological approach

The methodology focuses on capturing participants’ subjective perspectives, their interactions, and processes of meaning attribution, which are subsequently analyzed through a rule-guided interpretive framework (11). Fundamental methodological principles include an iterative process alternating between data collection and analysis, and the application of open, axial, and selective coding procedures as well as considering multiple interpretations and perspectives (process of “understanding others”) (14). To promote heterogeneity of perspectives, the research team strategically maximizes contrast across three key dimensions: theoretical knowledge, professional backgrounds, and levels of involvement in exercise planning and execution.

This principle of heterogeneity not only shapes the composition of the analysis team but also guides the broader research approach, which deliberately incorporates reflexivity regarding the researchers’ roles throughout the investigative process and integrates this awareness into the analytical framework. The epistemological foundation of the researchers’ orientations aligns with constructivist principles, drawing particularly on theoretical developments from second-generation GTM scholars (15). This perspective informs the entire research process, from study design through data interpretation, ensuring consistency with contemporary qualitative research paradigms in social science inquiry.

### Data collection

#### General Measures

Independently of the present research project, the training courses and exercises are planned by the external service provider Institut für Gefahrenabwehr (Institute for Hazard Prevention, Institut für Gefahrenabwehr GmbH, Zum Ehrenmal 12, 53809 Ruppichteroth, Germany, represented by Hanns Roesberg, M.Sc.) in close coordination with the Crisis and Disaster Management Unit of Heidelberg University Hospital.

Each participant will attend one of two available training and exercise sessions. The training sessions are scheduled for April 29, 2025, and May 20, 2025, while the exercise sessions will take place on May 26 and May 27, 2025.

The primary aim of the training is to prepare the hospital incident command system at Heidelberg University Hospital to effectively handle crisis situations. A continuous evaluation process (including, but not limited to pre-briefing, de-briefing, observation, feedback, coaching.) therefore takes place as part of the training courses and exercise scenarios to achieve the stated objectives. Participant observation is part of this evaluation process. As described by Jorgensen (16), it emphasizes understanding interactions from the participants’ perspectives within a flexible, evolving research process. It involves qualitative, case-oriented inquiry, relationship-building in the field, and the use of direct observation alongside other methods. Data are recorded through detailed field notes. To maintain an exploratory approach, predefined observation guidelines are avoided (17). Instead, sensitizing concepts (18) provide orientation, guiding attention to relevant phenomena without limiting openness.

Based on an empirical study of communication structures in staff work and the resulting findings on the effectiveness and efficiency of situation briefings (3), the aim is to raise awareness of the following characteristics in particular: the communication of different role expectations, awareness of the targeted use of language to achieve action goals, the professional and social familiarity of staff members, the structured design of the situation briefing, the clear definition of the group of participants and the functionality of technical communication systems and their acceptance.

#### Study specific Measures

Data systematically collected as part of the exercise on May 26 and 27—such as observation sheets, field notes, and photographic documentation of flipcharts—will be secondarily used as an additional source of information for the scientific evaluation of the exercise. This applies to all documents related to the content of the staff framework exercise.

All employees who agreed to participate in the study and who gave written informed consent to participate in a semi-structured interview will be contacted personally and an individual appointment will be scheduled with the interviewer.

To conduct the interviews, the project study team developed a semi-structured interview guide in accordance with the S^2^PS^2^ (collect, sort, check, delete, subsume) procedure (19). The guideline is divided into an open introduction with the aim of prompting a narrative and is supplemented by probe questions in order to provide alignment in structure (see Table 2).

**Table 2:** Interview Guide for the OrAKEL Study.

| Table 2: Interview Guide for the OrAKEL Study |  |
| --- | --- |
| **Introduction** | – Purpose of the interview<br>– Assurance of confidentiality and data protection<br>– Option to decline answering questions |
| **Demographic Information** | – Age, gender<br>– Professional experience and duration in current position<br>– Prior experience in disaster preparedness exercises or real-life emergencies |
| **Understanding Professional Context & Exercise Roles** | – What role did you assume during the exercise?<br>– Did this role feel realistic? Why (or why not)? |
| **Phase: Unfreeze**<br>**Checkpoints:**<br>✓ Initial crisis awareness (irritation, diffusion)<br>✓ Role transition (conscious vs. unconscious)<br>✓ Self-reflection on adaptability<br>✓ Relevance of role changes (rituals) | **(E) How did you perceive the situation when notified of the hospital's IT emergency?**<br>(P) How did you experience bearing responsibility for crisis management in the Hospital Emergency Management Team?<br>(P) As a leader/team member, how did you perceive the shift from routine operations to emergency mode? |
| **Phase: Change**<br>**Checkpoints:**<br>✓ Communication under temporary leadership<br>✓ Reflection on leadership approach<br>✓ Unexpected challenges in implementation | **(E) How was communication structured during the emergency?**<br>(P) How did you perceive team communication and collaboration?<br>(P) Who primarily organized or influenced communication?<br>(P) Did communication evolve during the scenario?<br>**(E) How did your tasks and role change during the emergency? Describe in detail.**<br>(P) How did you experience this shift?<br>(P) Which challenges or changes were most impactful?<br>(P) Did you consciously deviate from protocol? What motivated this?<br>**(E) How were leadership responsibilities reassigned?**<br>(P) Did you adjust your leadership style? If so, how?<br>(P) Which roles or individuals were most critical? Why?<br>**(E) How was crisis communication managed?**<br>(P) Were there misunderstandings or uncertainties? How were they resolved?<br>(P) What could have improved communication? |
| **Phase: Refreeze**<br>**Checkpoints:**<br>✓ Return to the regular communication structure<br>✓ Evaluation of the outcomes and reflection on the exceptional situation | **(E) How did you experience the transition back to normal operations?**<br>(P) What was particularly noticeable or challenging?<br>(P) How would you describe the post-crisis adjustment in communication and leadership?<br>(P) Did crisis management alter your perspective on daily functions?<br>**(E) How do you personally assess the success of crisis resolution?**<br>(P) What were the three main problems encountered?<br>(P) Which measures/decisions were most effective? |
| **Conclusion** | – Should any insights be shared internally or with other hospitals/organizations?<br>– In one sentence: What is your takeaway from this exercise?<br>– If you could make one improvement to crisis response—what would it be? |

(E = Exploratory question with the goal of prompting a narrative; P = Probe questions to provide alignment in structure)

The interview guide will be progressively refined through an iterative process of data collection and analysis. Following the completion of 3–5 initial interviews, the study team will re-evaluate the guide and make detailed adjustments as needed. Subsequently, in line with the principle of theoretical sampling, additional interviews will be conducted until theoretical saturation is achieved.The entire iterative procedure will be documented in accordance with the quality criteria of transparency and reproducibility.

The interviews will be conducted in accordance with the predefined study guidelines. At the start of each interview, participants will receive a comprehensive overview of the study’s objectives and procedures, including details on its duration, implementation, and the data collection and evaluation process.

The interviews are usually conducted web-based using video conferencing software on the basis of existing licenses (Microsoft Teams or Webex). Once all interviews have been conducted, data collection for the study is complete. A pseudonymized transcription of the interviews is carried out using MAXQDA software (VERBI Software, 1989-2024). In the subsequent evaluation phase, all available data, i.e. transcripts of the participant observation, documents created during the exercise, protocols, evaluation by the external service provider, and the transcripts of the interviews will be evaluated as part of an analysis group.

#### Ethics and Dissemination

The names of the employees and all other confidential information collected as part of the study are subject to confidentiality, the provisions of the German Federal Data Protection Act (BDSG) and the State Data Protection Act (LDSG) of Baden-Württemberg, Germany as well as those of the General Data Protection Regulation (GDPR). The study will be conducted in accordance with the Declaration of Helsinki and the current version of the Professional Code of Conduct for Physicians of the Baden-Württemberg Medical Association. The study was approved by the Ethics Committee of the Medical Faculty of Heidelberg University (S-065/2025) and was registered on ClinicalTrials.gov (DV 100 as a Framework for the Hospital Incident Command System (OrAKEL), Registration NCT06913010) prior to enrolment. In case of changes to eligibility criteria, outcomes, analyses or to relevant parties (e.g., investigators, REC/IRBs, trial participants, trial registries, journals, regulators) an amendment will be submitted to the Local Ethics Committee. It is planned to publish the data obtained during the study in a peer-reviewed scientific journal in accordance with the applicable data protection guidelines. Authors should adhere to the requirements of the ICMJE criteria (International Committee of Medical Journal Editors). The use of professional writers is not intended.

#### Perspective

To date, empirical research on the organizational structure of hospital incident command systems remains limited. While staff-based command models—adapted from military, fire, and police operations—are widely recommended in emergency planning frameworks (e.g., *Hospital Emergency and Response Planning Handbook*, 2020), their applicability and effectiveness in hospital settings have yet to be thoroughly examined. This study seeks to address this gap by providing empirical insights into the implementation of a staff-based hospital incident command system, with a specific focus on transition dynamics and internal communication processes.

## Acknowledgements

Not applicable.

## Author Contributions

MvdF, MR, JK, contributed to the conception and design. Drafting the manuscript: MvdF, MR, JK; Critically revision of the manuscript: MvdF, MR, JK, KB; EP; HS. All authors have read and agreed to the published version of the manuscript.

## Data availability

The datasets generated and analysed during the current study are not publicly available due local data protection regulation, but are available from the corresponding author on reasonable request.

## Literature

1. Handbuch Krankenhausalarm-und –einsatzplanung. Bundesamt für Bevölkerungsschutz und Katastrophenhilfe (BBK); 2020.

2. FwDV 100 Feuerwehr-Dienstvorschrift 100. Beschlossene Fassung des AFKzV https://www.lfs-bw.de/themen/gesetzevorschriften/fwdv/; 1999.

3. Celikler JM. Kommunizieren und Organisieren in komplexen Einsatzlagen. Wiesbaden: Springer Fachmedien Wiesbaden; 2023.

4. Hofinger G, Heimann R. Handbuch Stabsarbeit. 2. Hofinger G, Heimann R, editors. Berlin, Heidelberg: Springer Berlin Heidelberg; 2022. 3–10 p.

5. Schulze C, Welker A, Kühn A, Schwertz R, Otto B, Moraldo L, et al. Public Health Leadership in a VUCA World Environment: Lessons Learned during the Regional Emergency Rollout of SARS-CoV-2 Vaccinations in Heidelberg, Germany, during the COVID-19 Pandemic. Vaccines (Basel). 2021 Aug 11;9(8):887.

6. Waring S, Moisi I, Barrett C, Gordts S. Identifying what components of full-scale emergency exercises improve disaster response learning: A rapid evidence assessment. International Journal of Disaster Risk Reduction. 2024 Apr;104:104390.

7. Williams J, Nocera M, Casteel C. The Effectiveness of Disaster Training for Health Care Workers: A Systematic Review. Ann Emerg Med. 2008 Sep;52(3):211-222.e2.

8. Hsu EB, Jenckes MW, Catlett CL, Robinson KA, Feuerstein C, Cosgrove SE, et al. Effectiveness of Hospital Staff Mass-Casualty Incident Training Methods: A Systematic Literature Review. Prehosp Disaster Med. 2004 Sep 28;19(3):191–9.

9. Gowing JR, Walker KN, Elmer SL, Cummings EA. Disaster Preparedness among Health Professionals and Support Staff: What is Effective? An Integrative Literature Review. Prehosp Disaster Med. 2017 Jun 16;32(3):321–8.

10. Chan AW, Tetzlaff JM, Altman DG, Laupacis A, Gøtzsche PC, Krleža-Jerić K, et al. SPIRIT 2013 Statement: Defining Standard Protocol Items for Clinical Trials. Ann Intern Med. 2013 Feb 5;158(3):200–7.

11. Charmaz K. Constructivist grounded theory. J Posit Psychol. 2017 May 4;12(3):299–300.

12. Mey G, Mruck K. Grounded-Theory-Methodologie. In: Handbuch Qualitative Forschung in der Psychologie. 2. Wiesbaden: Springer Fachmedien Wiesbaden; 2020. p. 513–35.

13. Hennink M, Kaiser BN. Sample sizes for saturation in qualitative research: A systematic review of empirical tests. Soc Sci Med. 2022 Jan;292:114523.

14. Mey G, Mruck K. Grounded-Theory-Methodologie. In: Handbuch Qualitative Forschung in der Psychologie. Wiesbaden: VS Verlag für Sozialwissenschaften; 2010. p. 614–26.

15. Charmaz K, Thornberg R. The pursuit of quality in grounded theory. Qual Res Psychol. 2021 Jul 3;18(3):305–27.

16. Jorgensen D. Participant Observation. 2455 Teller Road, Thousand Oaks California 91320 United States of America : SAGE Publications, Inc.; 1989.

17. Flick U. Qualitative Sozialforschung: Eine Einführung. 10. König B, editor. Reinbek bei Hamburg: Rowohlt Taschenbuch; 2021.

18. Blumer H. What is Wrong with Social Theory? Am Sociol Rev. 1954 Feb;19(1):3.

19. Kruse J, Schmieder C, Weber KM, Dresing T, Pehl T. Qualitative Interviewforschung : ein integrativer Ansatz. 2. Weinheim; Basel: Beltz Juventa; 2015.

